# Frequency and spectrum of mutations in human sperm measured using duplex sequencing correlate with trio-based *de novo* mutation analyses

**DOI:** 10.1101/2024.02.13.24302689

**Authors:** Jonatan Axelsson, Danielle LeBlanc, Habiballah Shojaeisaadi, Matthew J Meier, Devon M. Fitzgerald, Daniela Nachmanson, Jedidiah Carlson, Alexandra Golubeva, Jake Higgins, Thomas Smith, Fang Yin Lo, Richard Pilsner, Andrew Williams, Jesse Salk, Francesco Marchetti, Carole Yauk

## Abstract

*De novo* mutations (DNMs) are drivers of genetic disorders. However, the study of DNMs is hampered by technological limitations preventing accurate quantification of ultra-rare mutations. Duplex Sequencing (DS) theoretically has < 1 error/billion base-pairs (bp). To determine the utility of DS to quantify and characterize DNMs, we analyzed DNA from blood and spermatozoa from six healthy, 18-year-old Swedish men using the TwinStrand DS mutagenesis panel (48 kb spanning 20 genic and intergenic loci). The mean single nucleotide variant mutation frequency (MF) was 1.2 x 10^-7^ per bp in blood and 2.5 x 10^-8^ per bp in sperm, with the most common base substitution being C>T. Blood MF and substitution spectrum were similar to those reported in blood cells with an orthogonal method. The sperm MF was in the same order of magnitude and had a strikingly similar spectrum to DNMs from publicly available whole genome sequencing data from human pedigrees (1.2 x 10^-8^ per bp). DS revealed much larger numbers of insertions and deletions in sperm over blood, driven by an abundance of putative extra-chromosomal circular DNAs. The study indicates the strong potential of DS to characterize human DNMs to inform factors that contribute to disease susceptibility and heritable genetic risks.

## Introduction

*De novo* mutations (DNMs) are a major cause of heritable genetic disorders including malformations, epilepsy, autism, cancer predisposition syndromes, and infertility.^1^ Globally, DNMs contribute to approximately 400,000 cases per year of developmental disorders.^2^ Indeed, a large part (30-70%) of genetic disorders have been attributed to DNMs^3–5^, accounting for a majority of inpatient pediatric admissions.^6,7^ Despite the clear importance of DNMs for individual and population health, there remain major gaps in our understanding of the endogenous and exogenous variables that mediate the rate, spectrum, and distribution of these mutations in the human genome.

Human pedigree studies have demonstrated that approximately 80% of DNMs have a paternal origin.^1,8,9^ Thus, male germ cells are the main source of mutations transmitted to children. There is very strong evidence that environmental factors significantly impact male germ cell mutation and DNM frequency in animal models.^10^ However, aside from ageing, the existence of human germ cell mutagens has yet to be convincingly demonstrated.^11^ Nonetheless, recent studies applying whole genome sequencing to human families provide intriguing support that certain pharmaceuticals and lifestyle factors cause DNMs in offspring *via* paternal germ cells.^10,12,13^

The major challenge to the study of spontaneous and induced mutagenesis is the lack of sufficiently accurate methodologies to measure the extremely low mutation frequencies occurring in the germ line (e.g., 1 x 10^-8^ per nucleotide in sperm^14^). The measurement of such low frequencies is hindered by the use of conventional next-generation sequencing (NGS) technologies that introduce errors in ∼ 1 of every 10^3^ nucleotides sequenced.^15^ Nonetheless, deep NGS has been applied to bulk sperm DNA to demonstrate that clonally appearing mosaic mutations occur and can be transmitted to the conceptus.^16^ Single cell sequencing of spermatozoa has been used to characterize mutations as well,^17^ but these technologies are also error-prone^18^ and may not be ideal for predicting DNM rates. Thus, more accurate NGS methodologies are required for the quantification of rare mutational events occurring during spermatogenesis to enable studies on the exogenous and endogenous drivers of DNMs.

Recently, advances in molecular barcoding paired with rigorous bioinformatic filtering approaches have been leveraged to produce error-corrected NGS methodologies that have an unprecedented level of sequencing accuracy.^15^ Duplex Sequencing (DS) eliminates errors by separately tagging, copying, and sequencing the top and bottom DNA strands from the same original duplex DNA molecule.^15^ Sequencing errors are removed by creating “families” of PCR duplicate reads originating from the same DNA molecule and building a duplex consensus sequence from the consensus sequence of the top and bottom strands. This improves the accuracy of NGS by 10,000 fold,^19^ to produce a theoretical error rate lower than 1 per billion nucleotides.^20^ Thus, these methods have great promise for quantification and characterization of both spontaneous and induced low-frequency mutations.^15^

Proof of principle studies show the potential utility of DS to identify rare mutations in both somatic tissues and germ cells. For example, DS was applied to quantify individual mutations down to a variant allele frequency (VAF) of 0.004% in blood to identify mutations that are of importance for drug-resistant relapse in acute lymphocytic leukemia.^21^ Applications in rodents suggest that DS can measure rare mutations (below 1 x 10^-7^ bp) induced by chemical exposures in various somatic tissues.^22,23^ A variation of DS (NanoSeq^14^) was applied to identify genome-wide mutations in sperm from a 21-year-old donor as a low mutation burden reference for somatic cell studies; the authors determined a mutation frequency (MF) of 1.8 x 10^-8^ bp, consistent with current estimates of the mutation rate in the paternal germline from trio studies (approximately 1.2 x 10^-8^ bp).^24^ Finally, Salazar et al.^25^ applied DS to the ∼ 4.5 kb coding region of the *FGFR3* gene, which is a mutational hotspot in the male germline, to reveal both unique and recurrent mutations, with an overall MF of approximately 6 × 10^−7^ across the locus. These initial studies show the promise of DS technologies for studying mutation burden in human sperm.

In the present study, we used DS to identify mutations in matched blood and sperm samples from six healthy, young men. We applied a 48 kb targeted DS approach comprising a panel of 20 genic and intergenic regions spread across the human autosomes.^26^ This hybrid capture panel provides an unbiased representation of sequence contexts within the human genome that includes loci with no known roles in cancer and no known selective advantages. The overarching aim was to investigate the suitability of DS to study sperm mutagenesis and infer DNM rates. Our results show concordance in the frequency and spectrum of DNMs measured by DS relative to whole genome sequencing of human pedigrees, supporting the power of this technology to quantify and characterize DNMs. We also demonstrate that DS detected putative extrachromosomal circular DNA (eccDNA) in high numbers in sperm (but not in blood), which could form the basis for further research.

## Materials and Methods

### Subjects, sample collection, and ethical approvals

Initially, 314 men from 17-20 years of age were recruited from the general Swedish population in 2008-2010 *via* the medical health examination at the National Service Administration in Sweden for possible military service.^27^ The men answered a questionnaire and provided blood and semen samples on the same day. After semen analysis, the remaining volume of the semen samples were frozen as raw semen. Blood was frozen down along with the semen at −80°C. For this study, we selected six 18-year-old men with normal sperm concentration^28^ and no chronic or long-term diseases.

The study was approved by the Ethical Review Board of Lund University and later by the Swedish Ethical Review Authority (approval number: 2021-02941). Health Canada’s participation in the bioinformatics analyses of DS data was approved by the Health Canada’s Research Ethics Board (approval number: REB 2022-004H).

### DNA extraction and Quality Control

Sperm DNA was extracted using a published method.^29^ Briefly, somatic cells in semen were removed by use of gradient centrifugation to avoid contamination of sperm DNA. Subsequently, sperm were homogenized with a Disruptor Genie (#SI-238l Scientific Industries, Bohemia, NY) in the presence of RLT buffer (Qiagen/Germany), Tris(2-carboxyethyl)phosphine (#77720; Pierce, Rockford, IL) and 0.2 mm steel beads (#SSB02; Next Advance, Averill Park, NY). DNA was extracted using the Qiagen QIAamp DNA micro kit (Cat. No. / ID: 56304, Qiagen, Germany). DNA from blood samples was extracted using the Qiagen DNeasy Blood and Tissue kit as described in the user’s manual (DNeasy Blood & Tissue Handbook 07/2020, Qiagen, Germany).

DNA quality was confirmed using an Agilent TapeStation (Agilent Technologies, Location); all samples passed quality control at TwinStrand Biosciences, Inc. (Seattle, USA) and had DNA integrity numbers of at least 6.6.

### Duplex Sequencing library preparation

DNA from sperm and blood were sent to TwinStrand Biosciences for library preparation and DS. The TwinStrand DuplexSeq™ Human Mutagenesis Assay (Human Mutagenesis Kit, TwinStrand Biosciences Inc., Seattle, WA, USA) with Enzymatic Fragmentation Module was used. The kit comprises 20 genomic loci, each 2.4 kb in size, spread across all autosomes but chromosomes 3 and 5 (Supplementary Table S1). Of these loci, ten are strictly genic, six are strictly intergenic, and four are mixed genic/intergenic. Sequencing libraries were prepared according to the manufacturer’s instructions. Briefly, 500⍰ng of DNA was used for enzymatic fragmentation, followed by end-repair, A-tailing, and ligation to DuplexSeq Adapters. Following an initial PCR amplification, the target regions were enriched using 120-nucleotide biotinylated oligonucleotide probes and subjected to bead-based capture, followed by another round of PCR, a second round of enrichment, and a final PCR. Prepared libraries were quantified, pooled, and sequenced on a NovaSeq 6000 (Illumina, San Diego CA, USA) at TwinStrand Biosciences.

### Bioinformatics – data processing and filtering

Raw sequencing data were processed and analyzed through the TwinStrand DuplexSeq™ Mutagenesis App (version 3.11.0) on DNAnexus®. The generation of consensus reads, post-processing and variant calling was performed as described previously.^22,23^ All statistical analyses were performed in R 4.2.0 and SPSS version 28, unless otherwise noted.

Further, a few additional filtering steps were implemented to ensure that only variants resulting from mutagenesis were considered. Identical mutations that appeared in more than one molecule in the same sample were considered to be derived from clonal expansion and were counted only once. The presence of no-calls (Ns) at the same position as a multi-nucleotide variant (MNV) in a read can generate false partial variant calls. Such instances are uncommon but become more likely at clonal or germline MNV sites. Therefore, single-nucleotide variants (SNVs) overlapping clonal MNVs (VAF ≥ 1%), were excluded from downstream analysis.

After observing clusters of SNVs near indel boundaries, we conducted a visual inspection of them which revealed no read-based support suggesting that they were introduced during the realignment process. Thus, we excluded all SNVs within 10 base pairs of indel boundaries, resulting in the removal of a total of 33 SNVs from our analysis. Intra-cohort contamination was defined as any mutation with VAF < 1% in one sample that also appears with a likely germline frequency (VAF > 30%) in at least one other sample. This led to the removal of 26 total variants across all samples, with one sperm DNA accounting for 16 of these. These two manual filters removed approximately 3% of somatic variant calls.

### Bioinformatics – mutation analysis

Only variants with a VAF ≤ 1% were included for mutation analysis since variants with a higher VAF were considered to be either (inherited) germline mutations or arising from mutations occurring during embryonic, fetal or possibly later development. Our initial dataset showed an unexpectedly high proportion of indels (insertions and deletions ≤ 1000 bp) and structural variants (SVs, insertions and deletions > 1000 bp, and all inversions) in the sperm DNA samples, and the indel length distribution did not match known distributions of germline indel length. Thus, we first focused our analysis exclusively on SNVs (i.e., excluded analysis of indels, structural variants and MNVs from both sperm and blood DNA).

The per sample MF was calculated by dividing the number of unique variants by the total number of duplex bases. We performed a correlation analysis (Pearson’s product-moment correlation [r]) between the MF in blood and sperm. In addition, we calculated MFs for each specific locus in the TwinStrand Human Mutagenesis panel and evaluated the Pearson correlation in the MF between sperm and blood by locus. We then analyzed whether there was a statistically significant difference in the MF between the ten genic and the six intergenic loci using Univariate Analysis of Variance (ANOVA). This analysis was conducted separately in blood and sperm.

The rate of SNV substitution subtype was calculated by dividing the number of each SNV subtype by the total duplex depth at all panel positions with the relevant reference allele. Accordingly, MF of C>A, C>G and C>T were divided by the total CG depth, whereas T>A, T>C and T>G were divided by the total TA depth. We plotted the MFs of the six types of base substitutions in blood and sperm and calculated the likelihood ratio statistic as described by Piegorsch and Bailer^30^ to determine whether the spectrum in blood differed from that in sperm. For C>T mutations, we also calculated the proportions occurring on CpG sites by looking at mutations occurring at NCG trinucleotides.

To identify the operational mutational signatures in the samples, we generated trinucleotide spectra by considering the flanking bases on either side of mutated bases ^31^. We used SigProfilerExtractor,^32,33^ ^34,35^, version 1.1.14, to decompose the trinucleotide spectrum observed in our data and assign them to known COSMIC mutational signatures (version 3.3) and reconstruct the original mutation matrix.^34^ We then calculated the cosine similarity between the spectra from both blood and sperm, and the COSMIC signatures. Finally, we calculated the cosine similarity between the spectra in blood and sperm.

Next, we compared the spectrum of single base substitutions in the blood with the spectrum and frequency of mutations detected in granulocytes and pediatric blood cells using two comparable methods, NanoSeq^14^ and EcoSeq, respectively^36^. To do this, we interpolated the proportions of the different types of substitutions from Figure 1d in Abascal et al. (2021)^14^ and from Figure S5 in the study by Ueda et al.^36^ We used the likelihood ratio statistic as above,^30^ to compare the spectrum in the blood of the six men and that of the granulocytes in the NanoSeq study^14^ and that of the blood cells of the EcoSeq study.^36^ We also compared the spectrum of SNV mutations in sperm DNA to the spectrum of DNMs measured in human pedigrees using whole genome sequencing. To do this, we collected raw data for 774,152 DNMs from eight published large trio cohorts^8,9,37–42^ and produced the mutation spectrum from these studies using the sigfit R package,^43^ again using the likelihood ratio statistic to see whether there was a statistically significant difference between the spectrum in sperm and the spectrum of the DNM.

**Figure 1.**
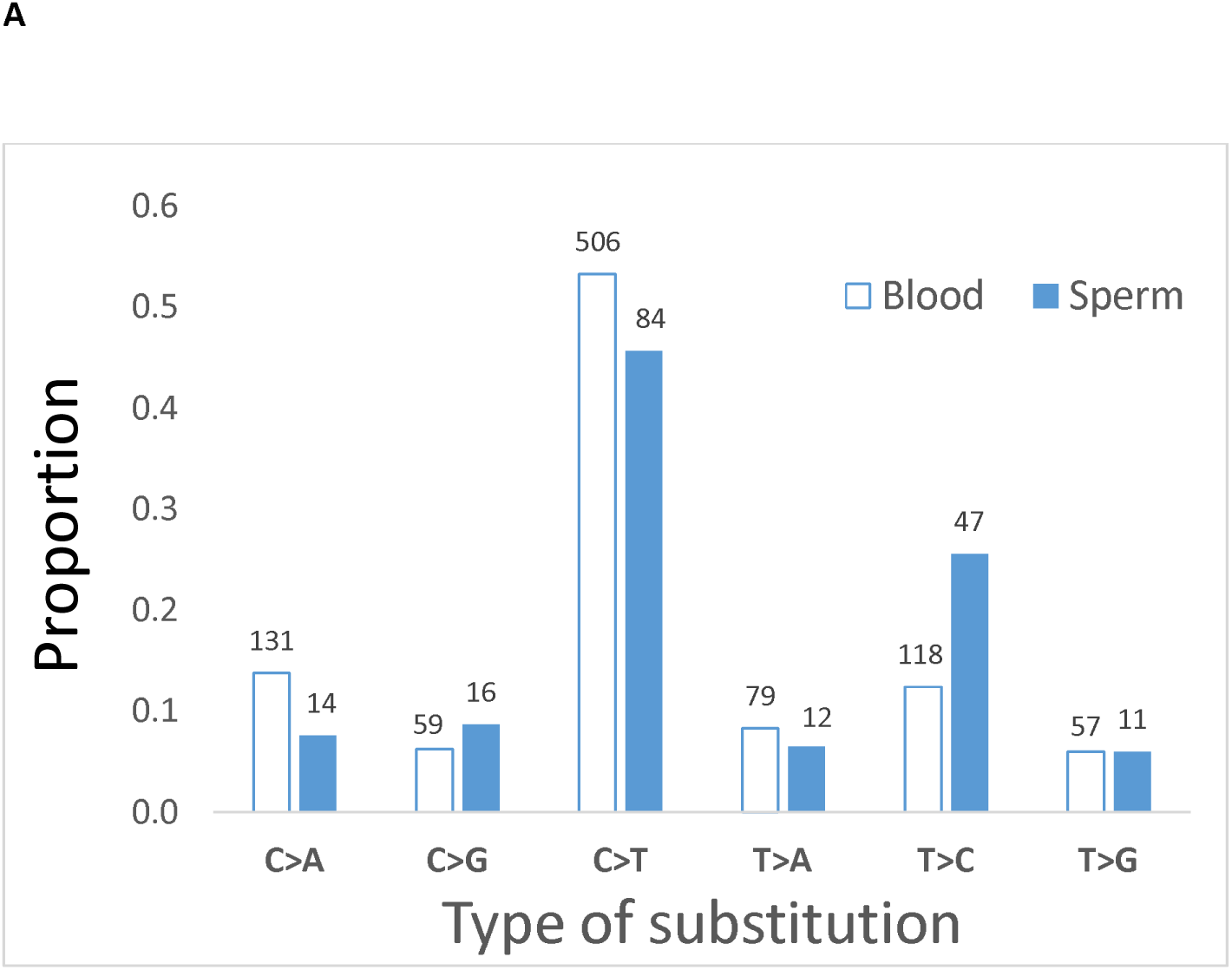
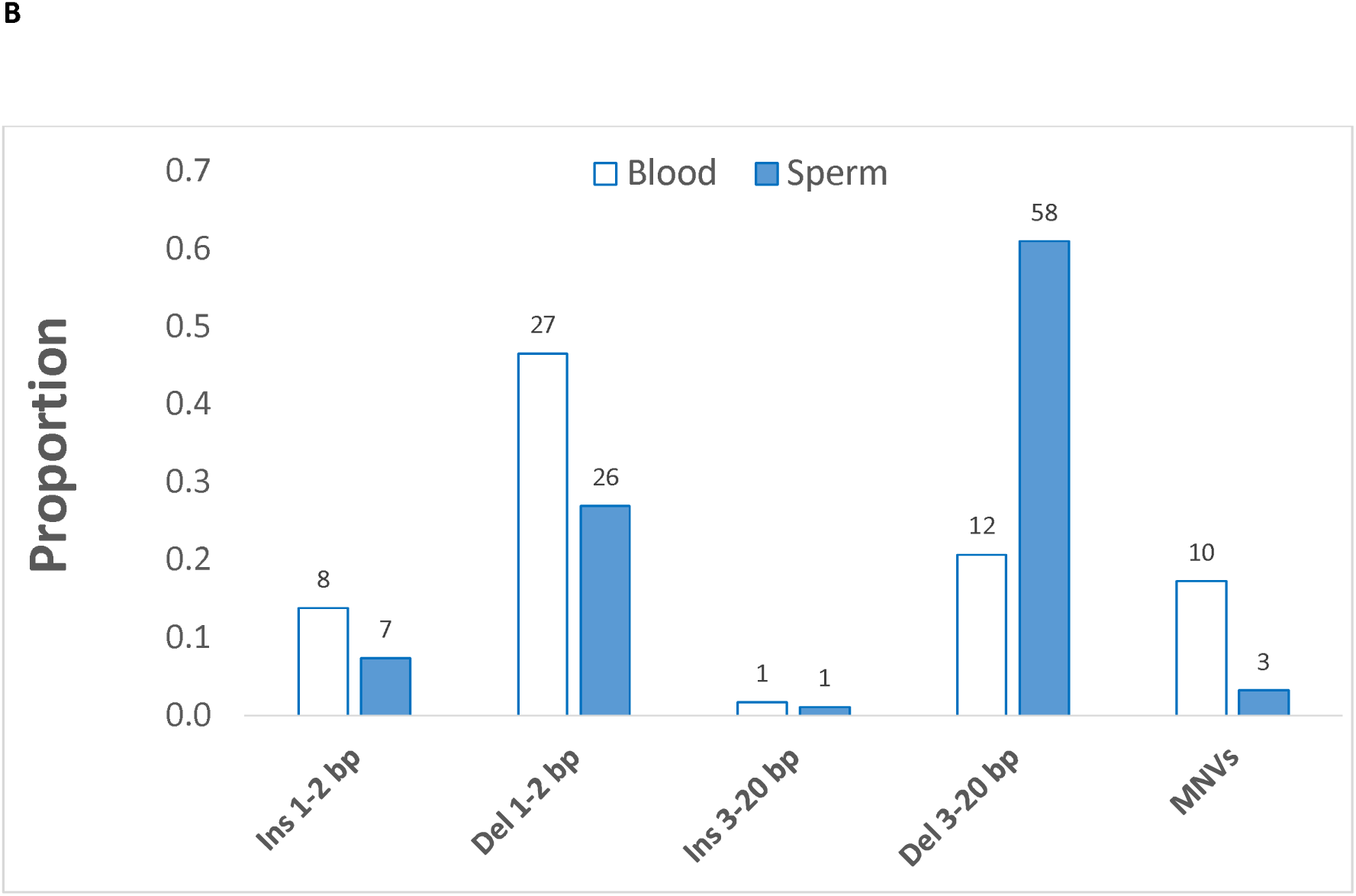
Proportions of single base substitutions (**A**) or short (≤ 20bp) insertions (Ins) and deletions (Del) as well as multi-nucleotide variants MNVs (**B**) in blood and sperm. The number of mutations for each type of substitution is shown above the bars. The spectrum of SNVs and the spectrum of the small insertions, deletions and MNVs differed between blood and sperm (both p < 0.001).

MNVs, indels, and SVs were analyzed separately, using R 4.1.2. Filtering, calculations, and data summarization were performed with dplyr and plots were generated with ggplot2 or Excel. First, indels were tabulated by size (1-2 bp, 3-20 bp, >20 bp) and identity (insertion or deletion). For complex variants involving the deletion of reference sequence combined with insertion of non-reference sequence, those with a net gain or loss in allele length were classified as insertions or deletions, respectively. This net change in allele length was defined as the indel length. We studied whether the spectrum of these type of insertions and deletions up to 20 bp as well as MNVs was statistically different between blood and sperm, using the likelihood ratio statistic as described above.^30^ Because the 1000 bp threshold between indels and SVs is an arbitrary parameter of variant calling, indels > 20 bp and SVs were then considered together and divided into categories based on type (insertion, deletion, inversion). Variant counts and frequencies were calculated for each size and type category and allele length was calculated as follows: (a) for variants originally called as indels, allele length was defined as the difference in string length between the reference and alternate alleles, as output by VardictJava; and (b) for all SV types, the allele length was calculated during variant calling (SVLEN field of the mut file).

### Bioinformatics - analysis of putative extrachromosomal DNA circles

Unlike for MF calculations, no VAF filter was applied during the analysis of large insertions. Because the distribution of allele lengths for large insertions was reminiscent of small extrachromosomal circular DNA (eccDNA), or microDNA,^44–48^ these variant calls were characterized further. First, the variant positions and alternate allele sequences were analyzed to determine whether they supported the type of junction expected for either circular DNA or chromosomal tandem duplications (junction fusing end and beginning of illustrative allele *ABCD*, subsequently referred to as a *D-A* junction). To identify *D-A* junctions, the alternate allele sequences were written to a FASTA file, which was used as input for matchPattern in BSgenome.Hsapiens.UCSC.hg38 (Bioconductor package), allowing 1 mismatch per 50 bp. The search was performed on a per-chromosome basis, only querying the chromosome containing the variant call. These search parameters yielded 0 or 1 match per sequence. Variants were considered to support a *D-A* junction if matchPattern returned a match located exactly 1 bp downstream of the variant position in the original mut file. Variant call records for SVs did not contain enough information to be classified with the matchPattern approach. Instead, duplex consensus reads supporting these variant calls were visualized in IGV to determine if they were consistent with *D-A* junctions, as described in detail below.

For fragment length analysis, duplex consensus reads supporting variant calls were visualized in IGV. First, the presence of a *D-A* junction was confirmed by inspecting supplementary alignments and/or BLAT-searching any soft-clipped sequences. For all manually inspected events (read pairs with a *D-A* junction), it was determined that all 5’ soft-clipped bases aligned at the other end of the allele, supporting the *D-A* junction, and any 3’ soft-clipped bases represented read-through into the DS adapter (only present for fragments < 142 bp in length). Most consensus read pairs containing *D-A* junctions aligned in discordant pairs, with one or both consensus reads having a substantial soft-clipped region with a supplementary alignment to the other end of the allele (spanning the *D-A* junction). For these read pairs, physical DNA fragment length was inferred by considering a pseudo-pair of reads made by the primary alignment of one read in the pair and the supplementary alignment of the other read in the pair and taking the difference between the right-most end of the reverse-aligned read and the left-most end of the forward-aligned read, including any 5’ soft-clipped bases. A few *D-A* junction-containing consensus read pairs aligned concordantly, in which case, the fragment length was inferred to be the computed insert size plus any 5’ soft-clipped bases. The distance between the 5’ ends of outward-facing read pairs (either primary-primary, or primary-supplementary) was noted based on visualization and confirmed to equal the allele length minus the inferred fragment length, except for the one confirmed chromosomal tandem duplication (TD).

To ensure a reasonable probability of detecting duplicated sequence if it was present, we selected the subset of apparent insertion variant calls with allele length less than the median insert size for all sperm DS libraries (233 bp) for systematic manual inspection. Considering the null hypothesis to be that all *D-A* junctions arose from chromosomal TDs, we expected that the length distribution of fragments supporting this subset of variant calls would be similar to the insert size distribution of the whole libraries, meaning that about half of fragments would be shorter than 233 bp, and the other half would be longer and that more than half of the fragments would be expected to include duplicated *ABCD* sequence (any fragment longer than its respective allele length). A binomial p-value was computed to test if the observed results were significantly different from the null hypothesis.

## Results

We used DS to measure MF and mutation spectra in blood and sperm from six healthy young males. We used blood as a reference somatic sample against which to compare the sperm baseline data, because research has already demonstrated effective performance of DS methodologies for identifying rare mutations in various somatic cell samples including blood.

Overall, an average of 440 million paired-end reads was produced per sample, with an average mean on target duplex error-corrected molecular depth of 27,045x, which generated approximately 1.2 billion total informative duplex bases per sample. In this analysis, we report unique mutations only; mutations appearing more than once in the same sample were assumed to have arisen from clonal expansion and were thus counted as one mutation, which may guard against over-estimating the MF.^49^ Of note, 96% of all SNV mutations (with a VAF < 1%) and 70% of non-SNV mutations were single count (non-clonal) mutations, respectively.

### Mutation frequencies in blood and sperm

We first established baseline MF in blood and sperm as measured by DS across the TwinStrand Human Mutagenesis panel. We then explored whether there was a correlation in MF between these tissues within men.

In blood, we identified 950 SNV mutations in the six samples from a total sequencing yield of approximately 7.9 billion duplex base pairs (Table 1). This resulted in a mean blood SNV MF of 1.2 x 10^-7^ (SD 0.19 x 10^-7^). SNV MFs per person varied from a low of 9.5 x 10^-8^ to a high of 1.4 x 10^-7^ in blood.

**Table 1.** Blood and sperm duplex sequencing depth, number of mutations* and mutation frequency (MF) for each participant.

| **BLOOD** |  |  |  |  |  |  |  |  |  |
| --- | --- | --- | --- | --- | --- | --- | --- | --- | --- |
| Participant | Duplex bases sequenced | # SNVs | SNV MF | # MNVs | MNV MF | # Indels | Indels MF | # SVs | SV MF |
| 1 | 1,354,242,708 | 137 | 1.0E-07 | 1 | 7.4E-10 | 8 | 5.9E-09 | 0 | 0 |
| 2 | 1,353,042,031 | 175 | 1.3E-07 | 2 | 1.5E-09 | 17 | 1.3E-08 | 0 | 0 |
| 3 | 1,200,540,488 | 114 | 9.5E-08 | 0 | 0 | 9 | 7.5E-09 | 1 | 8.3E-10 |
| 4 | 1,336,908,675 | 182 | 1.4E-07 | 2 | 1.5E-09 | 6 | 4.5E-09 | 0 | 0 |
| 5 | 1,341,009,611 | 188 | 1.4E-07 | 5 | 3.7E-09 | 11 | 8.2E-09 | 1 | 7.5E-10 |
| 6 | 1,310,508,887 | 154 | 1.2E-07 | 0 | 0 | 9 | 6.9E-09 | 1 | 7.6E-10 |
| *TOTAL* | *7,896,252,400* | *950* | *1.2E-07* | *10* | *1.3E-09* | *60* | *7.6E-09* | *3* | *3.8E-10* |
| **SPERM** |  |  |  |  |  |  |  |  |  |
| Participant | Duplex bases sequenced | # SNVs | SNV MF | # MNVs | MNV MF | # Indels | Indels MF | # SVs | SV MF |
| 1 | 1,129,894,519 | 32 | 2.8E-08 | 0 | 0 | 107 | 9.5E-08 | 13 | 1.2E-08 |
| 2 | 1,302,208,129 | 32 | 2.5E-08 | 1 | 7.7E-10 | 69 | 5.3E-08 | 4 | 3.1E-09 |
| 3 | 1,214,503,210 | 29 | 2.4E-08 | 0 | 0 | 67 | 5.5E-08 | 15 | 1.2E-08 |
| 4 | 1,301,231,218 | 33 | 2.5E-08 | 2 | 1.5E-09 | 105 | 8.1E-08 | 7 | 5.4E-09 |
| 5 | 1,236,182,547 | 31 | 2.5E-08 | 0 | 0 | 92 | 7.4E-08 | 10 | 8.1E-09 |
| 6 | 1,280,782,498 | 27 | 2.1E-08 | 0 | 0 | 98 | 7.7E-08 | 19 | 1.5E-08 |
| *TOTAL* | *7,464,802,121* | *184* | *2.5E-08* | *3* | *4.0E-10* | *538* | *7.2E-08* | *68* | *9.1E-09* |

\*single nucleotide variants (SNV); insertion/deletions (Indels, $\leq 1000$ bp); multi-nucleotide variants (MNV); and structural variants (SV, insertions and deletions $> 1000$ bp, and all inversions).

The SNV MF was nearly an order of magnitude lower in sperm. There were 184 SNV mutations across the six sperm samples, from a total yield of around 7.5 billion duplex base pairs. The mean SNV MF in sperm was 2.5 x 10^-8^ (SD 0.23 x 10^-8^), with a range of 2.1 to 2.8 x 10^-8^.

Thus, there was a 47% and 33% difference between the individuals with the highest and lowest SNV MF within blood and sperm, respectively. However, there was no correlation (Pearson’s r) between the SNV MF in sperm and that in the blood of the six men (r = −0.099, p = 0.85).

Similarly, the incidence of MNVs was higher in blood than in sperm (1.3 x 10^-9^ vs 4.0 x 10^-10^ per bp). However, there was a much lower frequency of insertions and deletions (indel) and structural variants (SV) in blood than in sperm. Specifically, we calculated an indel MF of 7.6 x 10^-9^ in blood versus 7.2 x 10^-8^ per bp in sperm, and an SV MF of 3.8 x 10^-10^ in blood versus 9.1 x 10^-9^ per bp in sperm (Table 1).

Taken together, these results show that both SNVs and MNVs were more common in blood than in sperm, whereas indels and SVs were more common in sperm than in blood.

### SNV mutation frequency analysis across the 20 independent loci

There was a high degree of inter-locus variability within both sperm and blood, with 5- and 6-fold differences between the highest and lowest locus-specific MFs in blood and sperm, respectively (Supplementary Tables S2 and S3). There was no correlation in locus-specific MF across the two cell types (Supplementary Figure S1). In addition, there was no statistically significant difference in the mean MF of genic versus intergenic loci (1.1 x 10^-7^ vs 1.4 x 10^-7^ [blood] and 2.5 x 10^-8^ vs 2.3 x 10^-8^ [sperm]).

### Mutation spectrum in blood and sperm

We explored whether DS in the two tissue types yields comparable single base substitution (SBS) mutation spectra.

In blood, the most common SNV was C>T with a mean MF of 1.5 x 10^-7^ (SD 0.22 x 10^-7^) constituting 53% of the six different SBS possible. This was followed by C>A [14% of SNV, MF 3.9 x 10^-8^ (SD 1.2 x 10^-8^)] and T>C [12% of SNV, MF of 2.6 x 10^-8^ (SD 0.64 x 10^-8^)] (Figure 1a, Supplementary Table S4). Similarly, C>T was the most common SNV in sperm with a MF of 2.6 x 10^-8^ (SD 0.53 x 10^-8^) bp, constituting 46% of the SBS (Figure 1a, Supplementary Table S5). However, unlike in blood, this was followed by T>C mutations in sperm with a frequency of 1.1 x 10^-8^ (SD 0.25 x10^-8^) bp (26% of the SBS).

The mutation spectrum in blood was significantly different than sperm (p < 0.0001). Overall, 35% and 36% of C>T mutations occurred at CpG sites in blood and sperm, respectively.

We also examined the spectrum of indels, first divided into insertions or deletions of 1-2 bp or 3-20 bp, and MNVs in blood and sperm (Figure 1b, Supplementary Table S6 and S7). Although the smallest category of indels occurred at similar frequencies in the two cell types, deletions of 3-20 bp were more frequent in sperm than in blood, with a statistically significant difference (p < 0.0001) between the two spectra (Figure 1b). Indels and SVs larger than 20 bp are summarized in Figure 2. These > 20 bp indels and SVs (deletions, insertions/duplications, and inversions) occurred at higher frequencies in sperm than in blood (Figure 2, Table S6 and S7). Deletions in sperm had a median length of 79 bp, with a maximum length of 2512 bp (Figure S2) and the insertions a multimodal size distribution (Figure S2).

**Figure 2.**
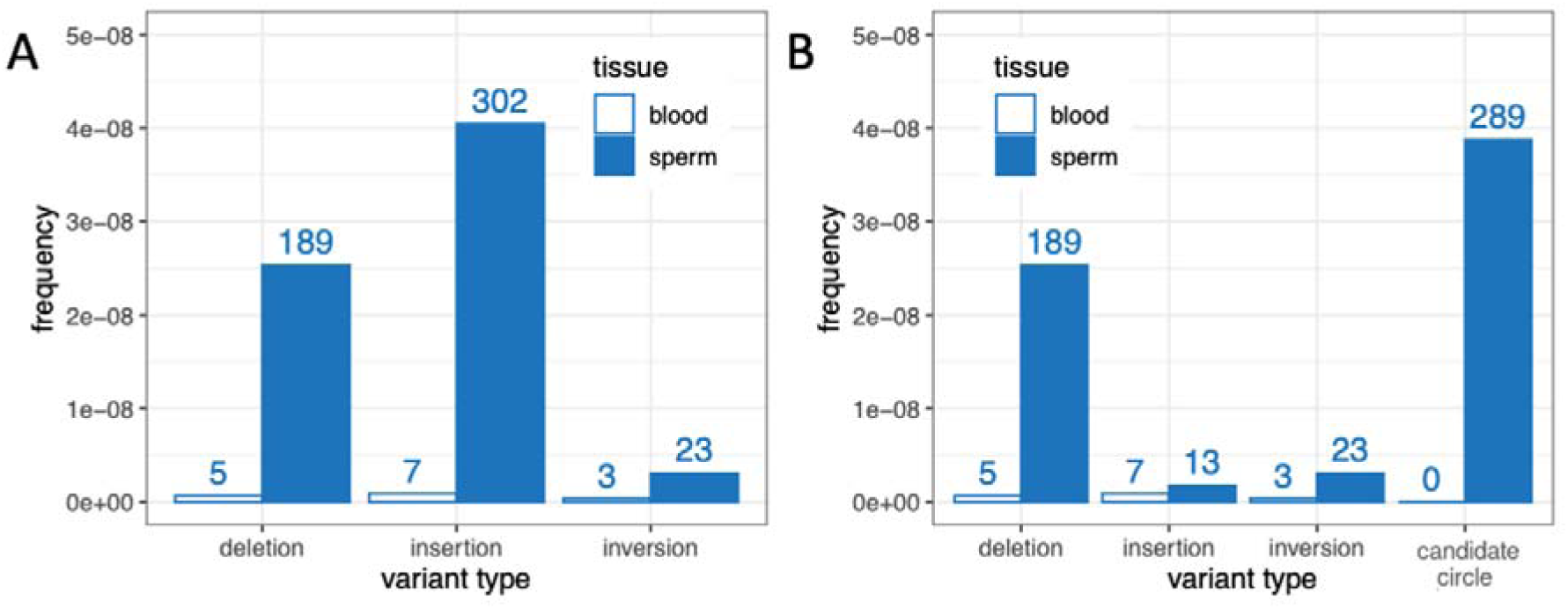
Frequencies of combined indels and SVs > 20 bp in length by tissue. For consistency with Table 1, only variants with VAF < 1% are included. **A)** Frequencies by type, as determined by the standard data analysis pipeline. **B)** Frequencies by amended type, with candidate circles plotted separately from insertions. Numbers above bars represent unique counts.

### Detection of putative circular DNA in sperm

The majority of apparent large insertions were called based on split alignment of a duplex consensus read to the end and beginning of a reference allele (Figure 3A i-iii). Given an illustrative reference allele, *ABCD*, this type of alignment suggests a junction that joins the end of the reference allele (*D*) to the beginning of the reference allele (*A*) and will be referred to as a *D-A* junction. *D-A* junctions can be formed by a chromosomal TD of *ABCD* (Figure 3A v) or when the *ABCD* allele is excised and circularized (Figure 3A iv). Figure 3B shows the periodic allele length distribution of apparent insertions, color-coded by whether or not they have a *D-A* junction. This analysis considers insertions of any VAF, whereas previous MF analyses were restricted to variants with VAF <1%. Across the six sperm samples, 96% of apparent insertions longer than 20 bp (308 out of 320) and 100% of apparent insertions longer than 125 bp (n = 308) contain *D-A* junctions (Figure 3B). No *D-A* junctions were detected in matched blood samples (n = 7 insertions > 20 bp). The length distribution of *D-A* junction-containing apparent insertions is strikingly similar to that reported for smaller extrachromosomal circular DNAs (eccDNA), also called microDNAs, in various cell types including sperm. _44-48_

**Figure 3.**
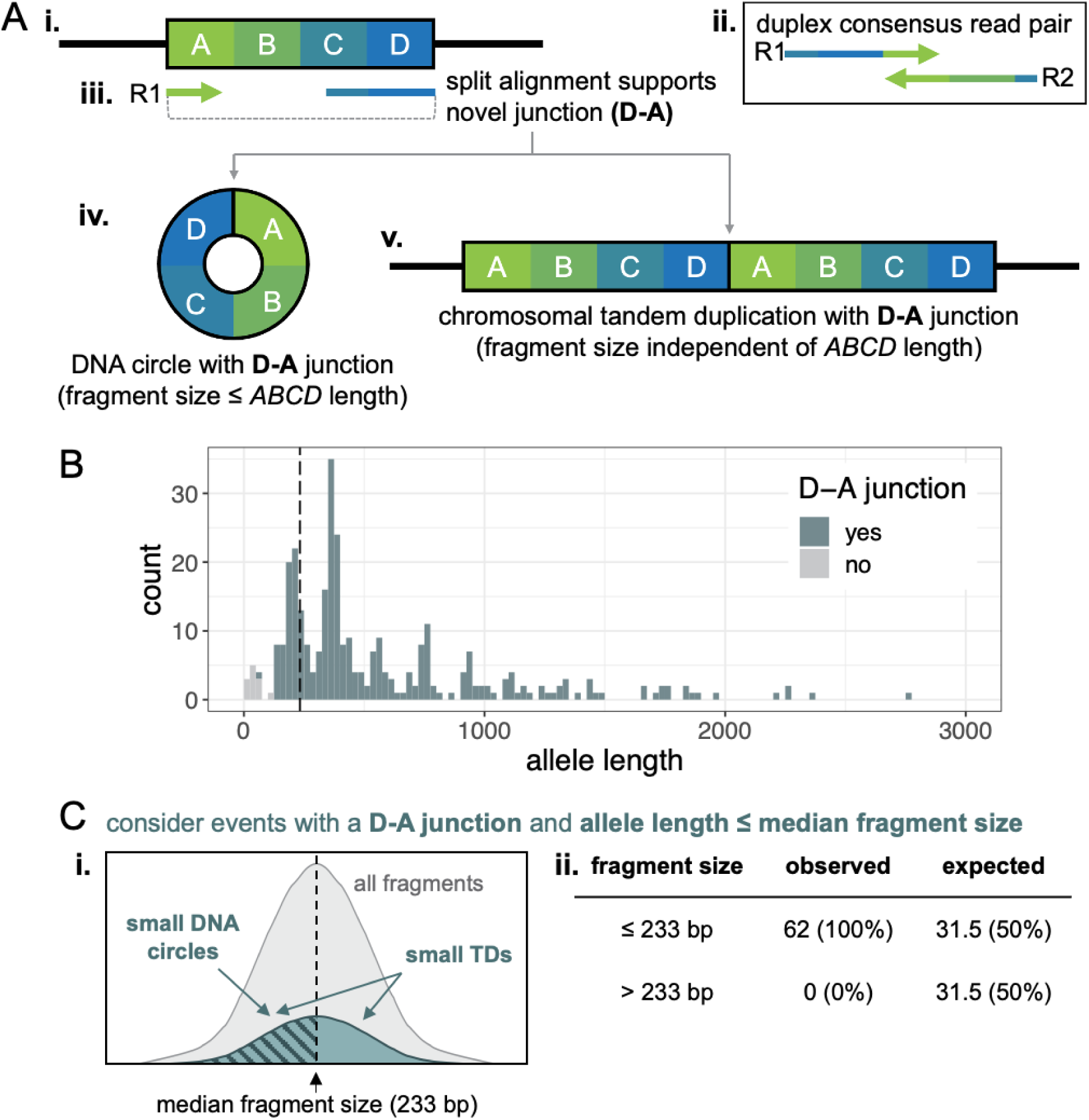
**A)** Schematic of **(i)** a reference allele *ABCD*, **(ii)** a duplex consensus read pair, **(iii)** split alignment of the R1 consensus read supporting a novel junction between the D and A (*D-A* junction), and two possible alternate alleles containing D-A junctions. Both **(iv)** excision and circularization of *ABCD* and **(v)** a chromosomal tandem duplication (TD) of *ABCD* create a D-A junction. **B)** Histogram of allele length for insertion calls ≥ 20 bp identified in sperm DNA, colored by whether they contain a D-A junction. The vertical dashed line is at the median fragment size of the libraries (233 bp). **C)** Considering a subset of events with allele length smaller than the median fragment size and containing a D-A junction (teal events left of vertical dashed line in B, n= 62), **(i)** chromosomal TDs should have fragment sizes that mirror the distribution of the whole library (teal and grey distributions, respectively), with fragment size independent of allele length. DNA circles in this subset should have fragment sizes less than or equal to the allele size, and thus, less than the median fragment size (diagonal hashes). **(ii)** All observed fragments were smaller than the median fragment size, significantly different from the expected distribution for chromosomal TDs (binomial p-value = 2.17 x 10^-19^).

To determine whether the *D-A* junctions detected by DS might arise from microDNAs, we explored the relationship between the lengths of DNA fragments containing *D-A* junctions and the length of *ABCD* alleles. Genomic DNA is enzymatically fragmented during DS library preparation and the size distribution of DNA fragments in final DS libraries can be approximated by the insert size distribution of the sequenced libraries (insert size is calculated as the distance between the reference genome alignment of the 5’ ends of read 1 and read 2 in a read pair). If *D-A* junctions arise from chromosomal TDs, the size distribution of the fragments containing the junction should be independent of the size of the duplicated *ABCD* allele and similar to that of the whole library (Figure S3). Additionally, any fragments larger than the *ABCD* allele will contain two copies of at least part of the *ABCD* sequence (e.g., *CDABCD*) and may also contain flanking non-*ABCD* sequence (Figure S3). Conversely, circular DNA molecules must be cleaved one or more times to generate linear DNA fragments and the resulting DNA fragments in the final library will be shorter than or equal to the length of the DNA circle, which is defined as the length of the *ABCD* allele (Figure S3).

To ensure a reasonable probability of detecting a duplicated sequence if present, for the following analysis we selected all *D-A* containing apparent insertions with an *ABCD* allele length of less than or equal to the median insert size of the six sperm libraries (233 bp, n=62) and manually reviewed the aligned duplex consensus reads supporting each call. If most or all *D-A* containing fragments in this subset arise from chromosomal TDs, about half of fragments would be larger than 233 bp and half would be smaller, as allele length and library fragment length should be independent (Figure 3C, entire teal distribution, Figure S3). Also, at least half of the events should have a fragment size larger than the *ABCD* allele length, and thus include at least some duplicated *ABCD* sequence and possibly non-*ABCD* flanking sequence (Figure S3). Conversely, all microDNA-derived fragments should be smaller than or equal to 233 bp, smaller than or equal to the *ABCD* allele length, and not contain any duplicated or flanking sequence (Figure 3C, portion of distribution with diagonal hashes; Figure S3). All 62 apparent insertions with allele length less than or equal to 233 bp had fragment lengths less than 233 bp, which is highly inconsistent with *D-A* junctions arising primarily from chromosomal TDs (binomial p-value = 2.17 x 10^-19^), and implies that most *D-A* junctions arise from putative microDNAs. Furthermore, 61 out of the 62 apparent insertions had fragment sizes shorter than their respective *ABCD* allele length, consistent with a microDNA origin. One chromosomal TD was identified conclusively: a 75 bp allele on a 180 bp fragment, containing duplicated and flanking sequence. Notably, this was the smallest *D-A* junction-containing variant call in the dataset and was much smaller than all other *D-A* junction-containing alleles (Figure 3B).

Based on these results, we consider all *D-A* junction-containing events with allele length greater than 125 bp to be candidate microDNAs. By this definition, we detected 307 candidate microDNAs across the six sperm samples (average 51 per sample, SD 18), yielding an average frequency of 4.1 x 10^-8^ (SD 1.4 x 10^-8^) in sperm. When only candidate circles with VAF <1% are considered, for consistency with other mutation frequency calculations, the average is 48 (SD 16) unique candidate microDNAs per sperm sample, or an average frequency of 3.9 x 10^-8^ (SD 1.3 x 10^-8^), which is higher than all other large indel (>20 bp) and SV types combined (Figure 2B). No candidate microDNAs were identified in the six matching blood samples (Figure 2B).

### Comparison of mutation spectra measured by Duplex Sequencing to published profiles and *de novo* mutations in children

We sought to further validate our results by comparing the frequencies and spectra determined by DS to other published studies. In addition, since our primary interest is in understanding DNMs in the offspring, we explored the predictivity of the measured sperm mutation spectrum to that inherited in the F1.

First, we examined the concordance of the MF and spectrum in blood samples with results of similar analyses obtained in granulocytes^14^ or blood^36^ to support the accuracy of our somatic cell mutation analysis. The estimated blood SNV MF herein was 1.2 x 10^-7^ (SD 0.19 x 10^-7^), which is similar to the reported MF of granulocytes identified by NanoSeq^14^ of 0.8 x 10^-7^, or of pediatric blood cells by EcoSeq (0.9 x 10^-7^).^36^ Moreover, highly similar mutation spectra were observed when comparing our data with that observed in granulocytes^14^ and pediatric blood cells,^36^ with no statistically significant difference between the spectrum in blood of the six men and in granulocytes (p = 0.86) and the pediatric blood cells (p = 0.49), all depicted in Figure 4.

**Figure 4.**
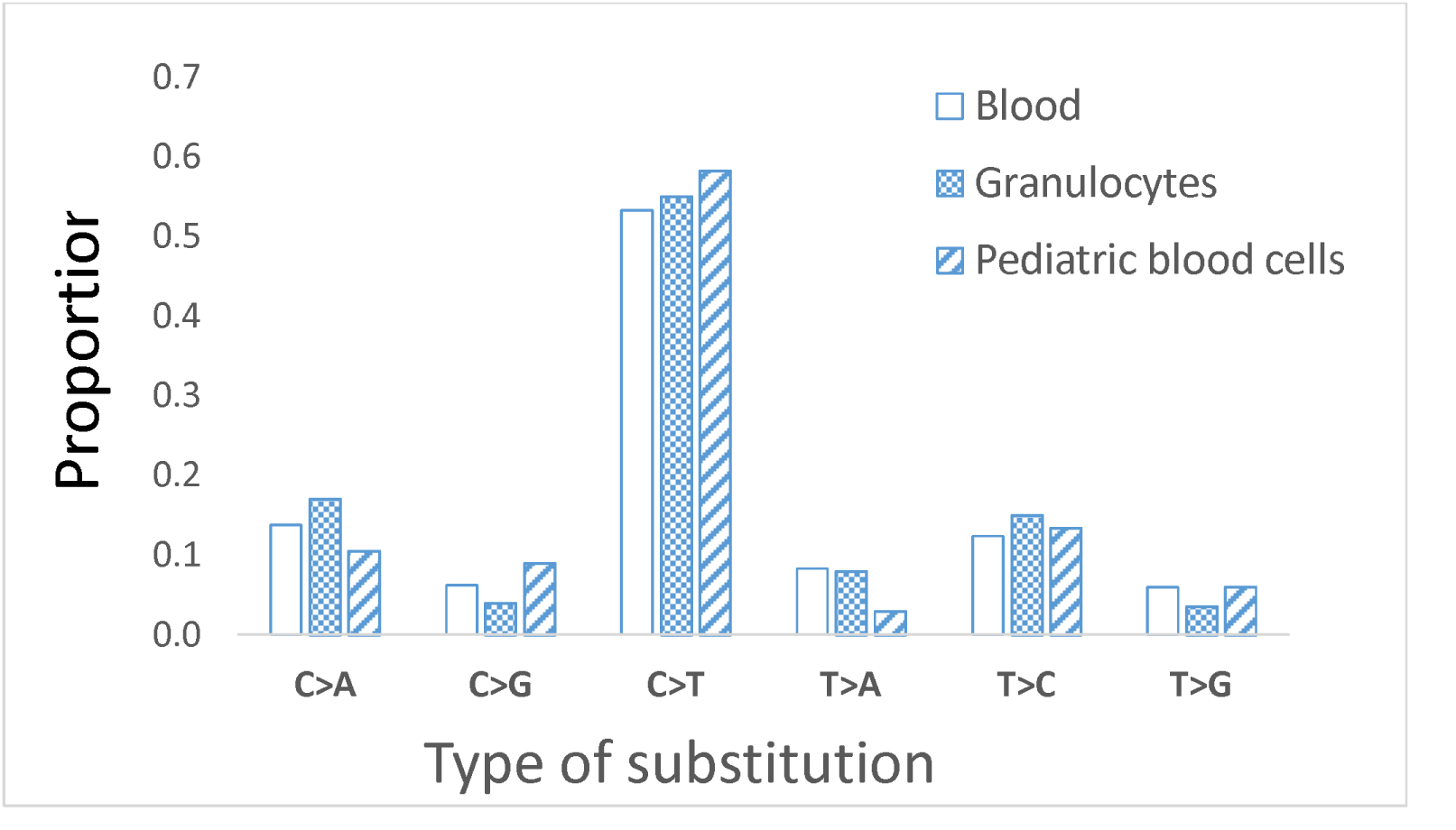
Proportions of single-base substitutions in the blood of the six men (present study) compared to proportions found in human granulocytes^14^ and pediatric blood cells,^36^ with no statistically significant difference between the spectrum in blood of our six men, and in the granulocytes (p = 0.86), and the pediatric blood cells (p = 0.49)

Next, we compared the sperm mutations identified by DS with > 774,000 human DNMs detected by NGS from eight large independent trio cohorts. ^8,9,37–42^ This is particularly important because the unique mutations measured by DS cannot be validated by targeted resequencing, whereas targeted resequencing was used to validate the DNMs in the pedigrees. The average DNM frequency across pedigree studies ^8,9,24,41,42^ is approximately 1.2 x 10^-8^, which is approximately 2-fold lower than our combined sperm SNV MF of 2.5 x 10^-8^. However, despite massive differences in the numbers of mutations comprising these data sets, we found a virtually identical mutation spectrum (p = 0.93 for difference) in the sperm compared with DNMs measured in offspring (Figure 5).

**Figure 5.**
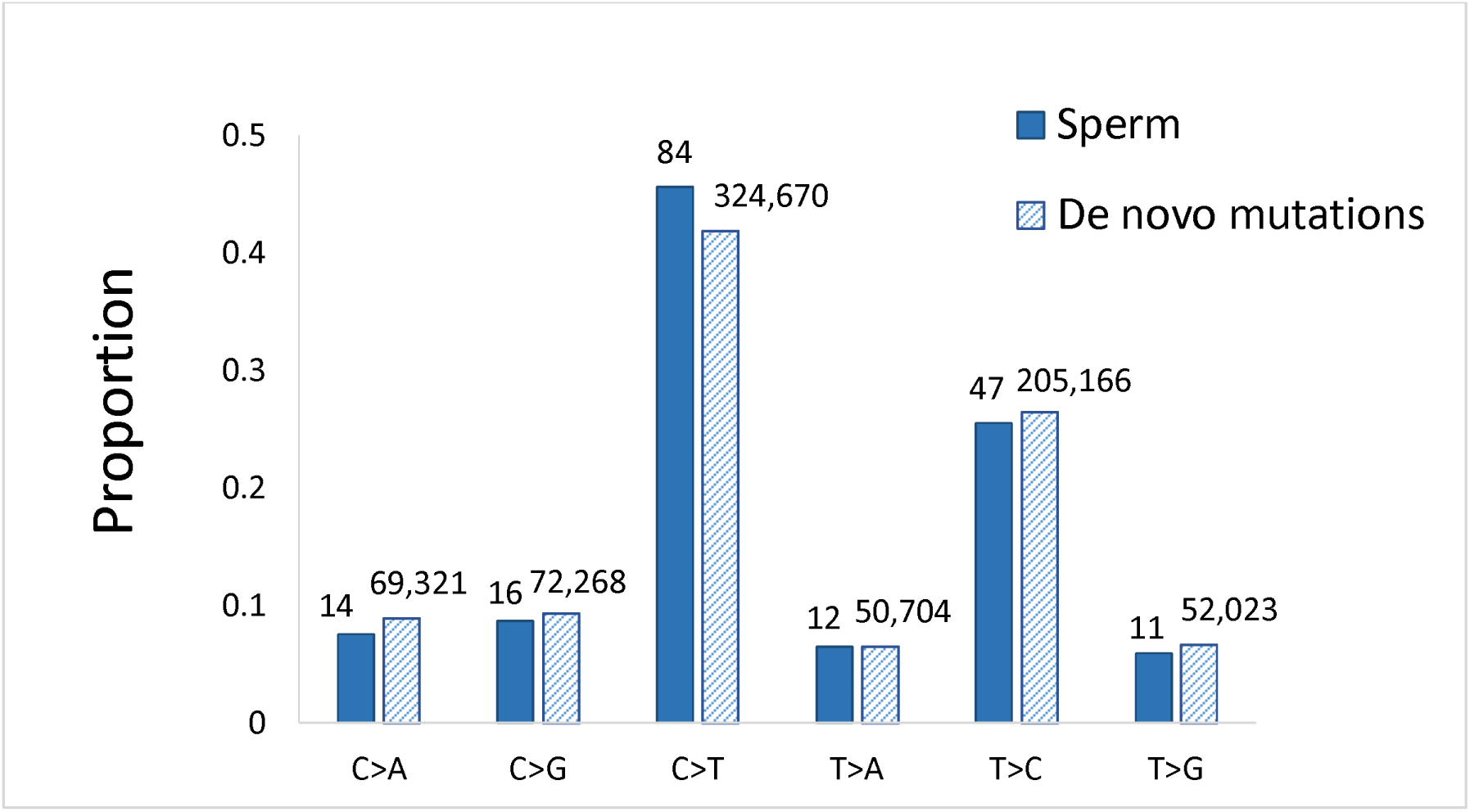
Proportion of different single base substitutions in sperm (present study) and *de novo* mutations identified in children from sequencing human pedigrees (p for difference = 0.93). Data on *de novo* mutations are compiled from previously published studies.^8,9,37–42^

We also analyzed the mutated bases in blood in the context of their adjacent flanking bases using SigProfilerExtractor. The generated trinucleotide spectrum matched a reconstructed signature with a combination of the following signatures (that may represent separate associated damage-or repair-related mechanisms^50^): 49% SBS40, 20% SBS1, 16% SBS11, and 15% SBS5, with a cosine similarity of 0.96 (Figure 6A). An identical analysis in sperm using the same tool, matched 88% to SBS5 and 12% to SBS1, with a cosine similarity of 0.87 (Figure 6B). The cosine similarity between the extracted signature in blood and sperm was 0.83.

**Figure 6.**
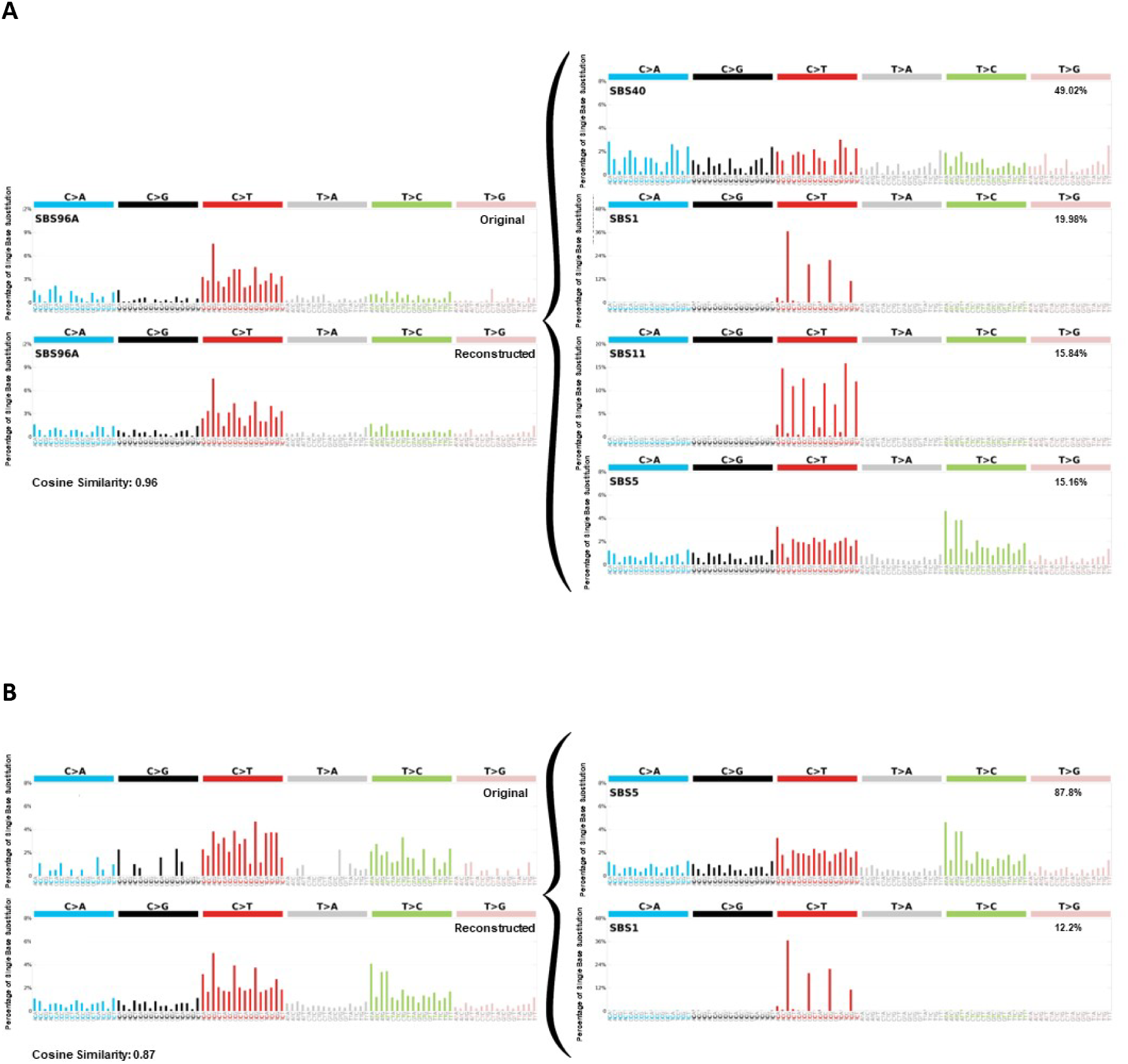
In both blood and sperm, the reconstructed^34^ mutational profiles with the highest similarity to the ones we found in blood and sperm were derived primarily from COSMIC mutational signatures associated with clock-like (i.e., age-dependent) processes. A) The trinucleotide mutational profiles from mutations in blood showed the highest cosine similarity to a combination of SBS40, SBS1, SBS11 and SBS5. B) The trinucleotide mutational profiles from mutations in sperm showed the highest cosine similarity to a combination of SBS5 and SBS1.

## Discussion

DS in six healthy 18-year-old men (with normal sperm profiles) revealed an SNV MF of 1.2 x 10^-7^ per bp in blood and 2.5 x 10^-8^ per bp in sperm. These MFs are within the expected range for these cell types. In addition, the SNV mutation spectra in blood were nearly identical to spectra from blood cells and pedigree analyses from previous studies, providing validation for the accuracy of the DS spectral results. These findings support that DS analysis of sperm DNA accurately models heritable mutagenesis. In contrast, we observed significant differences in mutation spectra between blood and sperm. Most notably, the sperm samples exhibited a much higher incidence of large indels and SVs. A detailed characterization revealed that these large indels/SVs constituted putative small eccDNAs that are present at high frequency in sperm but virtually absent in blood. Overall, we show that DS can be used to quantify intriguing and distinct features of mutagenesis in human sperm, providing an unprecedented opportunity to study the drivers of paternally-mediated heritable DNMs.

As expected,^51^ the SNV MF in blood was higher than in sperm. The MF in blood in our study was within the range of previously published data on pediatric blood cells,^36^ granulocytes,^14^ and other somatic cell organoids.^52^ Sperm SNV MF was in the same order of magnitude as DNM rates measured by whole genome sequencing of pedigrees (around 1.2 x 10^-8^).^8,41,53,54^ There was no correlation in the individual-specific SNV MF between blood and sperm across the men. We caution that this is a very small sample size for this analysis. Previous studies have shown higher and concordant MFs in the blood and sperm of men with defective DNA polymerase proofreading, supporting a potential correlation between MFs across tissues.^55^ However, polymorphisms in DNA polymerase and/or repair genes are expected to be strong genetic drivers of mutagenesis. The more subtle endogenous and exogenous factors leading to differences in MF within the men in our cohort may have much smaller effect sizes, requiring a larger sample size to observe correlations of MF across tissues. More research with a much larger sample size is clearly warranted to explore this relationship.

Our data revealed a relatively small amount of inter-individual variability in SNV MF within tissues in our six men. In fact, MF varied by only 1.3- (sperm) to 1.5-fold (blood) across the men. This is in-line with a 1.3-fold range in MF observed in cord blood across individuals using NanoSeq.^14^ Given that individuals in our study were all Caucasian men of European descent within a very narrow age range, and with no known genetic polymorphisms in DNA repair or replication genes, we expected inter-individual variability to be relatively low. Our results are also consistent with Blokzijl et al. (2016);^52^ these authors examined MF in clones of adult stem cells derived from primary colon, small intestine, and liver across a diverse population (with individuals spanning 3-87 years of age) and concluded that inter-human variability in MF was surprisingly low within tissues. Taken together, the low inter-individual variability observed with DS (and similar methods) supports its use as a method to precisely quantify MF.

In contrast to the relatively low degree of inter-human variability, we noted a larger extent of inter-locus variability. Indeed, SNV MF across loci varied by 5-fold in blood and 6-fold in sperm, although we caution that many fewer mutations per locus support the sperm data as compared to blood. Interestingly, there was no apparent correlation in the SNV MF of the loci between blood and sperm. Specifically, the most mutated loci in blood were not among the most mutated in sperm, and vice versa. In addition, we did not observe a significant difference between the intergenic and genic loci in either blood or sperm. These findings are at variance with prior studies showing a significantly higher MF in intergenic than genic regions for somatic mutations in human samples,^52,56^ as well as in somatic cells of mice^23^. The factors that modulate these observed locus-specific differences in mutagenic susceptibility across tissues remain to be identified.

The SNV mutation spectra were different between the blood and sperm of the six men (Figure 1a). The blood spectrum closely approximated that observed in human granulocytes,^14^ pediatric blood cells^36^ and other human cell types.^52^ The similarity with granulocytes is expected because they constitute the major type of leukocyte and cell type in blood that contains a nucleus, lending support to the accuracy of our results. The high degree of similarity with different technologies across various blood-based cell types provides weight of evidence supporting the accuracy of the acquired results.

We hypothesized that if DS provides an accurate method for SNV detection, we should see similar characteristics between the SNV spectra in sperm and DNMs in offspring. Remarkably, the DS SNV mutational spectrum was virtually identical to the SNV spectrum of DNMs in the aggregated data from pedigrees. The analysis revealed C>T mutations as the most dominant substitution, which is likely related to deamination of both unmethylated^57^ and methylated cytosines^58^, and which is the dominant mutation subtype in mammals.^59,60^ We found that 35% of C>T mutations in blood and 36% of those in sperm occurred at CpG sites, which is similar to DNMs in pedigrees, with proportions of C>T mutations occurring at CpG sites varying between 28% and 45%.^8,9,38,42^ Overall, our results suggest that the study of SNV mutations in human sperm by DS can be used to infer potential heritable risks.

Analyses of the trinucleotide spectra in our samples showed that they differed in terms of COSMIC mutational signatures, between blood and sperm. In fact, the spectra decomposed to SBS40, SBS1, SBS11 and SBS5 in blood, and to SBS5 and SBS1 in sperm. We note that SBS40, the most common SBS in the blood of our men, is highly similar to SBS5.^50,51^ SBS40 (like SBS5^61^) has an unknown etiology but is related to age.^50^ If we combined SBS40 and SBS5 into a single signature, as previously done by others to account for their similarity,^51^ then SBS5/40 and SBS1 were the most common signatures in both blood and sperm (Figure 6), with SBS5/40 being the most frequent. This also seems to be in line with the rather high cosine similarity of 0.83 between the trinucleotide spectra in blood and sperm (despite the difference found in the SNV spectra [Figure 1a]). Indeed, SBS5/40 and SBS1 have been reported as the most common signatures in 29 different cell types.^51^ Interestingly, SBS5 seems to be the signature that explains most of the intrinsic genetic variation in many different species.^61^ SBS5 and SBS1 were also among the three most common signatures in another study in sperm where DS was used to sequence the FGFR3 gene coding region.^25^ The proportions of the two signatures in sperm in our study (88% SBS5 and 12% SBS1) are also consistent with a study in seminiferous tubules^51^ and a study on DNMs in more than 20,000 families^12^, both of which reported 85% of SBS5/40 and 15% of SBS1. SBS5 is dominated by C>T and T>C mutations^62^, whereas SBS1 is characterized as deamination of 5-methylcytosine to thymine (https://cancer.sanger.ac.uk/signatures/sbs/). Both are considered clock-like signatures and correlate with the age of the individual. SBS11 has been associated with exposure to alkylating agents^63,64^ which is difficult to interpret in the context of sources of the blood samples herein (healthy young men). Taken together, the trinucleotide spectra we found are largely consistent with other studies of both somatic cells and germ cells.

Finally, in exploring mutation spectra we discovered a fascinating enrichment of putative eccDNA in sperm samples. Small eccDNAs, often called microDNAs, have been identified in many cell and tissue types in diverse eukaryotes, including plants, birds, rodents, and humans.^65–67^ The origin of these small circular DNA molecules is unclear, but most proposed mechanisms involve aberrant repair of DNA breaks.^65–67^ Potential functions of microDNAs are similarly elusive.^65–67^ Despite these unknowns, there is growing interest in microDNAs as biomarkers for an array of disease states.^65–67^ In addition, this is an important line of inquiry as it has been suggested that germline microDNAs could drive genome plasticity and evolution by integrating into chromosomal loci.^68^

Previous studies have identified microDNAs in human and rodent sperm using exonuclease-enrichment followed by high-throughput sequencing and/or by electron microscopy.^45,68,69^ One study, focusing on larger eccDNAs in human sperm, proposed that circular DNA forms during meiosis in regions depleted for meiotic crossovers.^69^ It is unclear if sperm microDNAs form by the same mechanism. Alternatively, sperm microDNAs could form during apoptosis, as demonstrated in some cell lines,^48^ with eccDNA formation seemingly linked with fragments that are protected by nucleosomes rather than bound to protamines.^70^ During sperm development, apoptosis can be triggered by failures of chromatin maturation in the testis or oxidative stress after spermiation.^71^ Sperm with apoptotically fragmented DNA are the primary source of DNA breaks measured as sperm DNA fragmentation and are associated with poor fertility outcomes.^71^ All subjects in the current study were young and healthy, with normal values of sperm DNA fragmentation (below 20%) measured with the Sperm Chromatin Structure Assay^72^ as part of a previous study.^73^ Further studies are needed to determine if sperm microDNA levels correlate with DNA fragmentation or other indicators of sperm quality and fertility.

More generally, we demonstrate that DS can detect candidate microDNAs without exonuclease enrichment, allowing for detection of mutations and putative microDNAs simultaneously. Because DS uses unique molecular identifiers to tag original DNA molecules prior to amplification,^20^ DS allows for more quantitative assessment of microDNAs than current methods that involve enrichment and/or rolling circle amplification. microDNA abundance increases when cells are exposed to diverse clastogens^46,74^ and pro-apoptotic compounds^46,48^, so we propose that their detection by DS-based mutagenesis assays, such as the one in this study, could provide a valuable read-out of genome instability. Future studies are needed to determine how well microDNA abundance correlates with standard clastogenicity measurements (e.g., comet and micronucleus assays) across a variety of cell and tissue types. The fact that microDNAs were not detected in blood is in line with the suggestion that microDNA is mostly derived from cells having undergone apoptosis-induced DNA fragmentation,^48^ a process that is well established to occur most abundantly in human sperm.^75^ However, it is also possible that the different DNA extraction methods used for the two sample types could have contributed to differences in microDNA detection.

In conclusion, our results indicate DS can reliably estimate very low frequencies of SNVs and characterize mutation spectra in human blood and sperm. The identified SNV mutation spectrum in human sperm was identical to that of inherited DNMs, supporting the high accuracy of these results. Intriguingly, our results suggest that DS detects the formation of eccDNAs, especially in sperm. The importance of these eccDNAs in relation to human inherited diseases warrants further scrutiny. Overall, our results support the great potential of DS as a powerful tool to identify factors contributing to germline mutagenesis and risk of DNMs in children.

## Data Availability

All data produced in the present study are available upon reasonable request to the authors

## Acknowledgements

We thank Professor Aleksander Giwercman for providing access to the herein used samples and research time to JA. We also thank all staff at the Reproductive Medicine Centre, Skåne University Hospital Malmö, Sweden, for help with recruitment of men, collection of samples and analysis of sperm chromatin integrity. We thank the Swedish Society of Medicine, ARMEC Lindebergs stiftelse, Maggie Stephens stiftelse, and the Faculty of Medicine, Lund University, for providing funding to make the study possible. FM acknowledges funding support from the Health Canada’s Genomic Research and Development Initiative. CLY acknowledges that this research was undertaken, in part, thanks to funding from the Canada Research Chairs Program (CRC award number CRC-2020-00060).

## Author contributions (CRediT author statement)

**Jonatan Axelsson**: Conceptualization, Resources, Data analysis, Writing - Original Draft, Funding Acquisition, Project administration.; **Danielle LeBlanc**: Formal analysis, Resources, Data Curation; **Habiballah Shojaeisaadi**: Formal analysis, Data Curation, Writing - Review & Editing; **Matthew Meier**: Formal analysis, Data Curation, Writing - Review & Editing; **Devon M. Fitzgerald:** Data analysis, Data Curation, Data Interpretation, Writing - Review & Editing; **Matthew Meier**: Formal analysis, Interpretation, Writing - Review & Editing; **Daniela Nachmanson:** Data Analysis, Data Curation Writing - Review & Editing; **Jedidiah Carlson:** Data Analysis; **Alexandra Golubeva:** Sample Processing, Data Curation; **Jake Higgins:** Data analysis, Data interpretation, Writing - Review & Editing; **Thomas Smith:** Data Analysis; **Fang Yin Lo:** Data Analysis; **Richard Pilsner:** Sperm sample processing, Writing - Review & Editing; **Andrew Williams:** Statistical analysis; **Jesse Salk:** Scientific advisor, Data interpretation; **Francesco Marchetti:** Supervision, Data interpretation, Writing - Original Draft, Review & Editing, Funding Acquisition; **Carole Yauk**: Supervision, Resources; Data interpretation, Writing - Original Draft Review & Editing, Funding Acquisition. All authors reviewed and approved the final version of the manuscript.

## Data availability statement

Data on DNA sequences can be found at NIH, National Library of Medicine through BioProject Accession: PRJNA928759

