## Supplementary for "Frequency and spectrum of mutations in human sperm measured using duplex sequencing correlate with trio-based *de novo* mutation analyses"

**Supplementary Table S1.** Human Mutagenesis Panel targets

| **Contig** | **Start** | **End** | **Description** | **Location relative to genes** | **Gene (Genome Data Viewer NCBI)** |
| --- | --- | --- | --- | --- | --- |
| chr1 | 84597127 | 84599527 | region_208 | Genic/ intergenic | LINC01461 |
| chr2 | 40162767 | 40165167 | region_2896 | Genic | SLC8A1-AS1 |
| chr4 | 22386244 | 22388644 | region_4173 | Genic/ intergenic | ADGRA3 |
| chr6 | 155239014 | 155241414 | region_5020 | Genic | TIAM2 |
| chr7 | 11732774 | 11735174 | region_5144 | Genic | THSD7A |
| chr8 | 51513056 | 51515456 | region_5520 | Genic | PXDNL |
| chr9 | 23709463 | 23711863 | region_5739 | Genic | ELAVL2 |
| chr10 | 128969037 | 128971437 | region_784 | Intergenic | |
| chr11 | 108510787 | 108513187 | region_1111 | Genic | EXPH5 |
| chr12 | 114115043 | 114117443 | region_1355 | Intergenic | |
| chr13 | 75803912 | 75806312 | region_1501 | Genic | LMO7 |
| chr14 | 74661755 | 74664155 | region_1725 | Genic | AREL1 |
| chr15 | 46089737 | 46092137 | region_1904 | Intergenic | |
| chr16 | 51754103 | 51756503 | region_2115 | Genic/ intergenic | LOC105371257 |
| chr17 | 70672726 | 70675126 | region_2378 | Intergenic | |
| chr18 | 5749264 | 5751664 | region_2457 | Genic | MIR3976HG |
| chr19 | 31831021 | 31833421 | region_2739 | Genic/ intergenic | LINC01837 |
| chr20 | 24153684 | 24156084 | region_3388 | Genic | LINC01721 |
| chr21 | 23665976 | 23668376 | region_3515 | Intergenic | |
| chr22 | 48262370 | 48264770 | region_3703 | Intergenic | |

**Supplementary Table S2.** Locations, number, and frequency of SNV mutations across the loci in the Duplex Sequencing Human Mutagenesis panel in *blood* from the six men.

| **Location,  Chromosome** | **Type** | **SNVs** | **Total duplex bases** | **SNV MF** |
| --- | --- | --- | --- | --- |
| 1 | Genic/intergenic | 21 | 4.1 x 10^8^ | 5.1 x 10^-8^ |
| 2 | Genic | 63 | 4.2 x 10^8^ | 1.5 x 10^-7^ |
| 4 | Genic/intergenic | 53 | 3.8 x 10^8^ | 1.4 x 10^-7^ |
| 6 | Genic | 30 | 4.3 x 10^8^ | 7.0 x 10^-8^ |
| 7 | Genic | 38 | 3.1 x 10^8^ | 1.2 x 10^-7^ |
| 8 | Genic | 62 | 3.9 x 10^8^ | 1.6 x 10^-7^ |
| 9 | Genic | 56 | 3.5 x 10^8^ | 1.6 x 10^-7^ |
| 10 | Intergenic | 60 | 4.0 x 10^8^ | 1.5 x 10^-7^ |
| 11 | Genic | 26 | 4.4 x 10^8^ | 6.0 x 10^-8^ |
| 12 | Intergenic | 74 | 4.4 x 10^8^ | 1.7 x 10^-7^ |
| 13 | Genic | 21 | 4.1 x 10^8^ | 5.1 x 10^-8^ |
| 14 | Genic | 14 | 4.3 x 10^8^ | 3.3 x 10^-8^ |
| 15 | Intergenic | 49 | 3.7 x 10^8^ | 1.3 x 10^-7^ |
| 16 | Genic/intergenic | 62 | 3.8 x 10^8^ | 1.6 x 10^-7^ |
| 17 | Intergenic | 41 | 3.7 x 10^8^ | 1.1 x 10^-7^ |
| 18 | Genic | 68 | 4.2 x 10^8^ | 1.6 x 10^-7^ |
| 19 | Genic/intergenic | 57 | 4.0 x 10^8^ | 1.4 x 10^-7^ |
| 20 | Genic | 48 | 4.3 x 10^8^ | 1.1 x 10^-7^ |
| 21 | Intergenic | 41 | 3.0 x 10^8^ | 1.4 x 10^-7^ |
| 22 | Intergenic | 66 | 4.3 x 10^8^ | 1.5 x 10^-7^ |
| *TOTAL* |  | *950* | *79 x 10^8^* | *1.2 x 10^-7^* |

**Supplementary Table S3.** Locations, number, and frequency of SNV mutations across the loci in the Duplex Sequencing Human Mutagenesis panel in the *sperm* DNA of the six men.

|  |  | **SNV mutations** | | |
| --- | --- | --- | --- | --- |
| **Location,**  **Chromosome** | **Type** | **SNVs** | **Total duplex bases** | **SNV MF** |
| 1 | Genic/intergenic | 11 | 3.8 x 10^8^ | 2.9 x 10^-8^ |
| 2 | Genic | 12 | 4.0 x 10^8^ | 3.0 x 10^-8^ |
| 4 | Genic/intergenic | 6 | 3.4 x 10^8^ | 1.8 x 10^-8^ |
| 6 | Genic | 14 | 4.1 x 10^8^ | 3.4 x 10^-8^ |
| 7 | Genic | 4 | 2.8 x 10^8^ | 1.4 x 10^-8^ |
| 8 | Genic | 14 | 3.6 x 10^8^ | 3.9 x 10^-8^ |
| 9 | Genic | 9 | 3.2 x 10^8^ | 2.8 x 10^-8^ |
| 10 | Intergenic | 16 | 3.9 x 10^8^ | 4.1 x 10^-8^ |
| 11 | Genic | 3 | 4.0 x 10^8^ | 7.0 x 10^-9^ |
| 12 | Intergenic | 6 | 4.4 x 10^8^ | 1.4 x 10^-8^ |
| 13 | Genic | 10 | 3.8 x 10^8^ | 2.6 x 10^-8^ |
| 14 | Genic | 13 | 4.2 x 10^8^ | 3.1 x 10^-8^ |
| 15 | Intergenic | 3 | 3.4 x 10^8^ | 8.7 x 10^-9^ |
| 16 | Genic/intergenic | 9 | 3.7 x 10^8^ | 2.5 x 10^-8^ |
| 17 | Intergenic | 7 | 3.5 x 10^8^ | 2.0 x 10^-8^ |
| 18 | Genic | 7 | 4.0 x 10^8^ | 1.8 x 10^-8^ |
| 19 | Genic/intergenic | 10 | 3.8 x 10^8^ | 2.7 x 10^-8^ |
| 20 | Genic | 10 | 3.8 x 10^8^ | 2.7 x 10^-8^ |
| 21 | Intergenic | 5 | 2.6 x 10^8^ | 1.9 x 10^-8^ |
| 22 | Intergenic | 14 | 4.4 x 10^8^ | 3.2 x 10^-8^ |
| TOTAL |  | 184 | 7.5 x 10^9^ | 2.5 x 10^-8^ |

**Supplementary Table S4.** Subtypes of SNV base substitutions in *blood*

| **Base substitution subtype** | **Numbers** | **Mutation Frequency Mean (per bp)** | **Standard deviation  (per bp)** | **Proportion (%)** |
| --- | --- | --- | --- | --- |
| **C>A** | 131 | 3.9 x 10^-8^ | 1.2 x 10^-8^ | 14% |
| **C>G** | 59 | 1.8 x 10^-8^ | 7.7 x 10^-9^ | 6.2% |
| **C>T** | 506 | 1.5 x 10^-7^ | 2.2 x 10^-8^ | 53% |
| **T>A** | 79 | 1.7 x 10^-8^ | 7.0 x 10^-9^ | 8.3% |
| **T>C** | 118 | 2.6 x 10^-8^ | 6.5 x 10^-9^ | 12% |
| **T>G** | 57 | 1.3 x 10^-8^ | 6.5 x 10^-9^ | 6.0% |
| ***Total*** | 950 | *NA* | *NA* | *100%* |

**Supplementary Table S5.** Subtypes of base substitutions in *sperm*

| **Base substitution subtype** | **Numbers** | **Mutation frequency Mean (per bp)** | **Standard deviation** | **Proportion (%)** |
| --- | --- | --- | --- | --- |
| **C>A** | 14 | 4.3 x 10^-9^ | 2.5 x 10^-9^ | 7.6% |
| **C>G** | 16 | 5.1 x 10^-9^ | 3.4 x 10^-9^ | 8.7% |
| **C>T** | 84 | 2.6 x 10^-8^ | 5.3 x 10^-9^ | 46% |
| **T>A** | 12 | 2.8 x 10^-9^ | 2.2 x 10^-9^ | 6.5% |
| **T>C** | 47 | 1.1 x 10^-8^ | 2.5 x 10^-9^ | 26% |
| **T>G** | 11 | 2.6 x 10^-9^ | 1.3 x 10^-9^ | 6.0% |
| ***Total*** | *184* | *NA* | *NA* | *100%* |

**Supplementary Table S6.** Subtypes, numbers, and MF of indels in *blood*

| **Indels** | | | | | | | | | | | | **MNVs** | | **SVs** | |
| --- | --- | --- | --- | --- | --- | --- | --- | --- | --- | --- | --- | --- | --- | --- | --- |
| **1-2 bp** | | | | **3-20 bp** | | | | **>20 bp** | | | |  | |  | |
| **Number** | | **MF** | | **Number** | | **MF** | | **Number** | | **MF** | | **Number** | **MF** | **Number** | **MF** |
| 35 | | 4.4 x 10^-9^ | | 18 | | 2.3 x 10^-9^ | | 12 | | 1.5 x 10^-9^ | | 10 | 1.3 x 10^-9^ | 3 | 3.8 x 10^-10^ |
| **Ins** | **Del** | **Ins** | **Del** | **Ins** | **Del** | **Ins** | **Del** | **Ins** | **Del** | **Ins** | **Del** |  |  |  |  |
| 8 | 27 | 1.0 x 10^-9^ | 3.4 x 10^-9^ | 1 | 12 | 1.3 x 10^-10^ | 1.5 x 10^-9^ | 7 | 5 | 8.9 x 10^-10^ | 6.3 x 10^-10^ |  |  |  |  |

Abbreviations: Del, deletions; Ins, insertions

**Supplementary Table S7.** Subtypes, numbers, and MF of indels in *sperm*

| **Indels** | | | | | | | | | | | | **MNVs** | | **SVs** | |
| --- | --- | --- | --- | --- | --- | --- | --- | --- | --- | --- | --- | --- | --- | --- | --- |
| **1-2 bp** | | | | **3-20 bp** | | | | **>20 bp** | | | |  | |  | |
| **Number** | | **MF** | | **Number** | | **MF** | | **Number** | | **MF** | | **Number** | **MF** | **Number** | **MF** |
| 33 | | 4.4 x 10^-9^ | | 59 | | 7.9 x 10^-9^ | | 446 | | 7.0 x 10^-8^ | | 3 | 4.0 x 10^-10^ | 68 | 9.1 x 10^-9^ |
| **Ins** | **Del** | **Ins** | **Del** | **Ins** | **Del** | **Ins** | **Del** | **Ins** | **Del** | **Ins** | **Del** |  |  |  |  |
| 7 | 26 | 9.4 x 10^-10^ | 3.5 x 10^-9^ | 1 | 58 | 1.3 x 10^-10^ | 7.8 x 10^-9^ | 262 | 184 | 3.5 x 10^-8^ | 2.5 x 10^-8^ |  |  |  |  |

Abbreviations: Del, deletions; Ins, insertions

Mutations in sperm per 108 bases

| Mutations in blood per 108 bases | MF in blood per 107 bases |
| --- | --- |
| 5.07643 | 2.90196 |
| 14.8804 | 4.12333 |
| 5.97926 | 0.747875 |
| 16.9848 | 1.35822 |
| 5.0905 | 2.63091 |
| 3.286 | 3.11499 |
| 13.3244 | 0.874368 |
| 16.3342 | 2.4505 |
| 11.1242 | 2.02638 |
| 16.3235 | 1.74842 |
| 14.3515 | 2.65971 |
| 14.8646 | 3.01555 |
| 11.2617 | 2.59036 |
| 13.8442 | 1.93391 |
| 15.3666 | 3.18981 |
| 13.8846 | 1.74631 |
| 6.97512 | 3.41632 |
| 12.2624 | 1.44057 |
| 15.7349 | 3.86008 |
| 15.9709 | 2.81775 |

**Supplementary Figure S1.** SNV mutation frequency in sperm vs in blood in the 20 different loci. R^2^ value derived from graph in Excel.

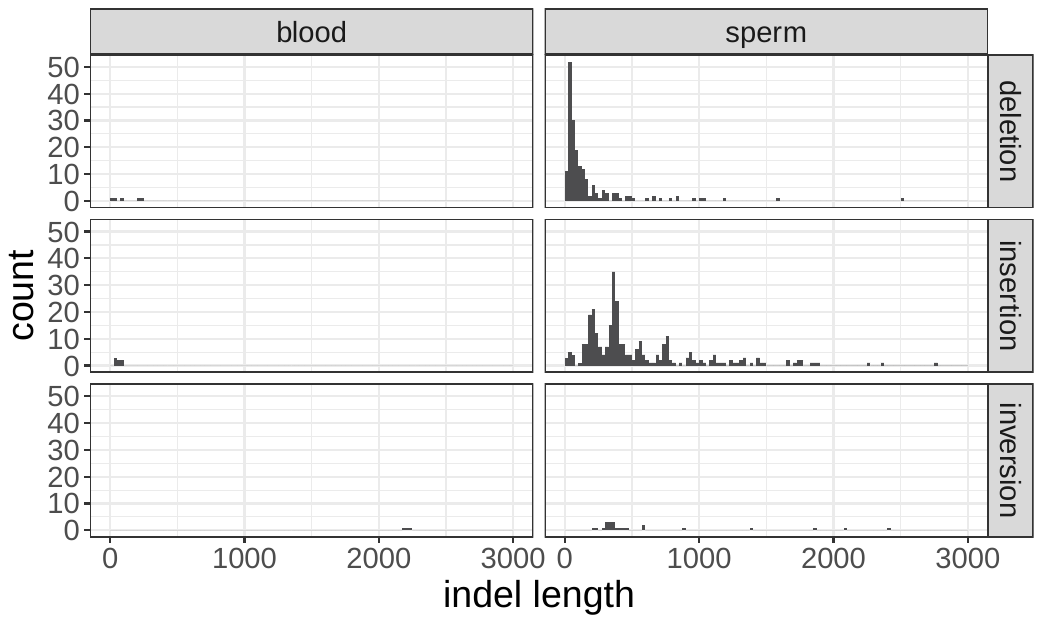

**Supplemental Figure S2.** Histograms of allele sizes for variants longer than 20 bp, with separate panels for tissues and variant types. For consistency with mutation frequency calculations, only variants with VAF < 1% are included.

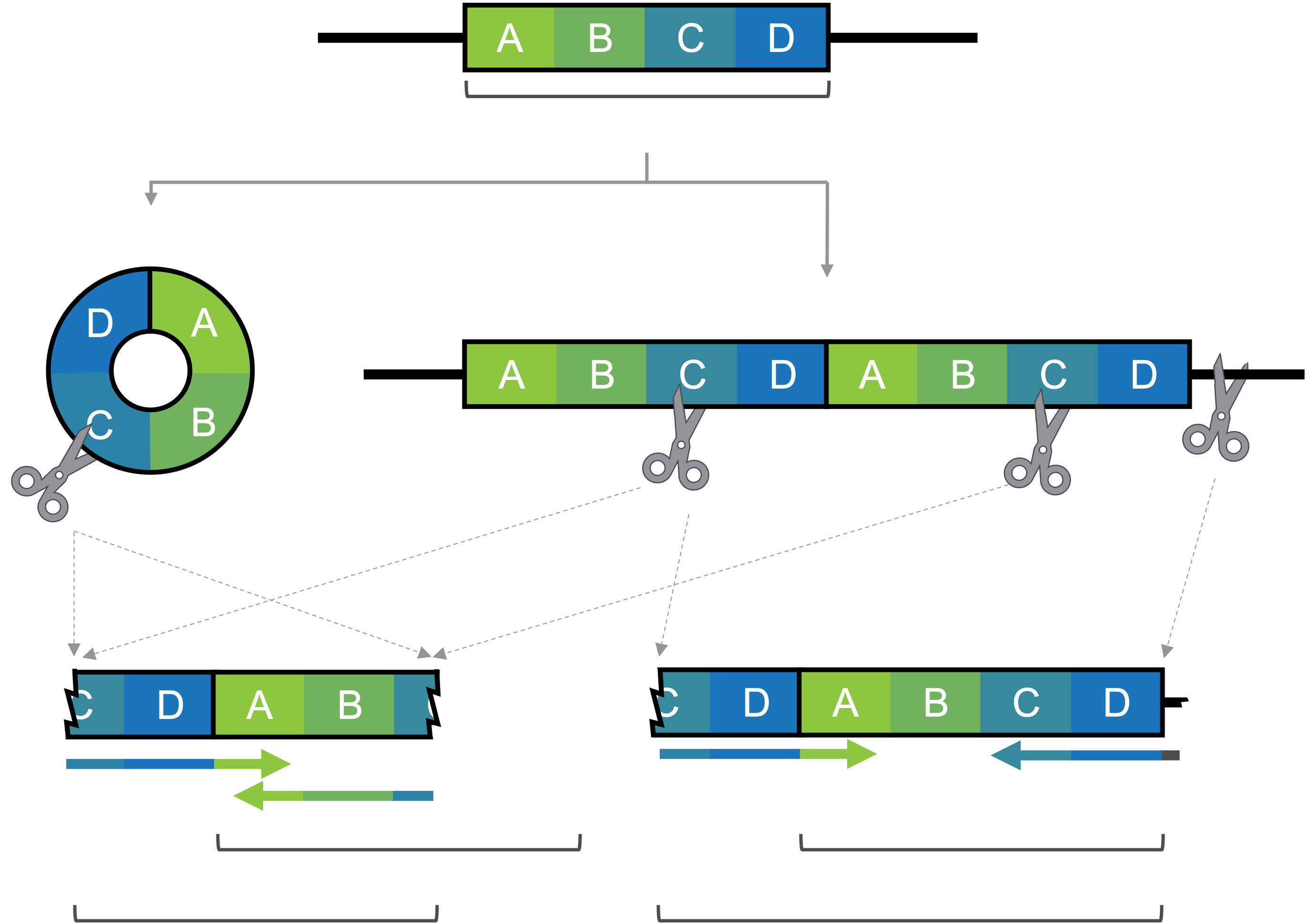

chromosomal tandem duplication with **D-A** junction

circular DNA with **D-A** junction

**from circle or TD**

(fragment size ≤ allele length, no duplicated sequence within fragment)

*allele length = 400 bp*

*allele length = 400 bp*

*fragment size ≤ 400 bp*

*fragment size > 400 bp*

**from TD only**

(fragment size > allele length**,** duplicated sequence within fragment)

*allele length = 400 bp*

B

C

A

**ii.**

**i.**

**ii.**

**i.**

**Supplemental Figure S3.** Schematic showing the relationship between allele length and fragment size for a D-A junction-containing molecule arising from circular DNA or a chromosomal tandem duplication*.* If a 400 bp long reference allele *ABCD* (A) forms a circular DNA (Bi), one cleavage by the fragmentation enzyme (scissors) will yield a linear fragment with length equal to the allele length of 400 bp (Ci). Two or more cleavages of the circular DNA molecule with yield fragments smaller than the allele length (not shown). A chromosomal TD of *ABCD* (Bii) can be cleaved into fragments smaller than (not shown), equal to (Ci), or larger than (Cii) the allele length of 400 bp. Note that the longer fragment (Cii) includes multiple copies of the reference allele sequences C and D.
